# Neural networks for endemic measles dynamics: comparative analysis and integration with mechanistic models

**DOI:** 10.1101/2024.05.28.24307979

**Authors:** Wyatt G. Madden, Wei Jin, Benjamin Lopman, Andreas Zufle, Benjamin Dalziel, C. Jessica E. Metcalf, Bryan T. Grenfell, Max S. Y. Lau

**Author notes:** Department of Biostatistics and Bioinformatics, Rollins School of Public Health, Emory University, Atlanta, Georgia, United States of America (Wyatt G. Madden, Max S. Y. Lau).

## Abstract

Measles is an important infectious disease system both for its burden on public health and as an opportunity for studying nonlinear spatio-temporal disease dynamics. Traditional mechanistic models often struggle to fully capture the complex nonlinear spatio-temporal dynamics inherent in measles outbreaks. In this paper, we first develop a high-dimensional feed-forward neural network model with spatial features (SFNN) to forecast endemic measles outbreaks and systematically compare its predictive power with that of a classical mechanistic model (TSIR). We illustrate the utility of our model using England and Wales measles data from 1944-1965. These data present multiple modeling challenges due to the interplay between metapopulations, seasonal trends, and nonlinear dynamics related to demographic changes. Our results show that, while the TSIR model yields more accurate very short-term (1 to 2 biweeks ahead) forecasts for highly populous cities, overall, our neural network model (SFNN) outperforms the TSIR in other forecasting windows. Furthermore, we show that our spatial-feature neural network model, without imposing mechanistic assumptions *a priori*, can uncover gravity-model-like spatial hierarchy of measles spread in which major cities play an important role in driving regional outbreaks. We then turn our attention to integrative approaches that combine mechanistic and machine learning models. Specifically, we investigate how the TSIR can be utilized to improve a state-of-the-art approach known as Physics-Informed-Neural-Networks (PINN) which explicitly combines compartmental models and neural networks. Our results show that the TSIR can facilitate the reconstruction of latent susceptible dynamics, improving both forecasts and parameter inference of measles dynamics within the PINN. In summary, our results show that appropriately designed neural network-based models can outperform traditional mechanistic models for short to long-term forecasts, while simultaneously providing mechanistic interpretability. Our work also provides valuable insights into more effectively integrating machine learning models with mechanistic models to enhance public health responses to measles and similar infectious disease systems.

**Author summary:** Mechanistic models have been foundational in developing an understanding of the transmission dynamics of infectious diseases including measles. In contrast to their mechanistic counterparts, machine learning techniques including neural networks have primarily focused on improving forecasting accuracy without explicitly inferring transmission dynamics. Effectively integrating these two modeling approaches remains a central challenge. In this paper, we first develop a high-dimensional neural network model to forecast spatiotemporal endemic measles outbreaks and systematically compare its predictive power with that of a classical mechanistic model (TSIR). We illustrate the utility of our model using a detailed dataset describing measles outbreaks in England and Wales from 1944-1965, one of the best-documented and most-studied nonlinear infectious disease systems. Our results show that, overall, our neural network model outperforms the TSIR in all forecasting windows. Furthermore, we show that our neural network model can uncover the mechanism of hierarchical spread of measles where major cities drive regional outbreaks. We then develop an integrative approach that explicitly and effectively combines mechanistic and machine learning models, improving simultaneously both forecasting and inference. In summary, our work offers valuable insights into the effective utilization of machine learning models, and integration with mechanistic models, for enhancing outbreak responses to measles and similar infectious disease systems.

## Introduction

Following the COVID-19 pandemic, there has been a marked increase in machine learning research focused on enhancing the forecasting of infectious diseases. This body of work primarily sought to develop highly predictive models for real-time application during the peak of the health crisis [1, 2]. A portion of these studies has endeavored to meld classical mechanistic approaches to infectious disease with machine learning, either through the post-hoc analysis of machine learning outputs in light of established disease dynamics [3, 4], or by directly integrating mechanistic insights into the machine learning models [5–7]. Our research advances these efforts by developing neural-network-based models tailored to the complex spatiotemporal multi-year transmission dynamics of endemic measles, leveraging a well-characterized infectious disease system and a rich historical dataset describing outbreaks in pre-vaccination England and Wales.

Measles is one of the most highly transmissible and strongly immunizing pathogens. Spatiotemporal patterns of pre- and post-vaccination measles incidence are among the most well-documented, and well-studied, nonlinear infectious disease systems. Measles exhibits complex spatiotemporal dynamics driven by the interplay between seasonal forcing, susceptible recruitment due to births and spatial coupling between populations. These dynamics range from regular multiannual infection patterns in large populations [8] to coexisting attractors [9]. By contrast, measles dynamics in small highly vaccinated populations dominated by chaotic patterns driven by stochastic extinction [10]. For example, before widespread vaccination in the late 1960s, measles epidemics in England and Wales were dominated by highly regular periodic (often biennial) cycles in large cities whose populations are at, or above, the Critical Community Size (CCS) *−* the population size required to maintain endemic transmission *−* of approximately 300,000 individuals [11]. Following the widespread vaccination in the late 1960s, the epidemics shifted from highly regular cycles to largely irregular dynamics [12]. Due to its simple natural history and long time series of data, measles incidence in England and Wales has provided a fruitful testing ground for better understanding spatiotemporal nonlinear epidemiological dynamics, and developing semi-mechanistic statistical modeling approaches more broadly [13–18].

A suite of previous analyses has demonstrated the utility of deterministic and stochastic (semi-) mechanistic models, notably the time-series-SIR (TSIR) model [13], a discrete approximation of the S-I-R model, and other successful inferential approaches including particle filtering [19], in characterizing the dynamics in large urban populations. However, in general, these models have not primarily focused on generating long-term forecasting accuracy. While machine learning models, including a recent work leveraging the Least Absolute Shrinkage and Selection Operator (LASSO) [15], have shown improved forecasting skills for endemic measles dynamics, they generally lack deep mechanistic interpretability. These models also do not explicitly consider spatial interactions between locations which is a known driver for measles transmission, particularly, between less populous locations (e.g., small towns) and population centers (e.g., core cities) [16]. To this end, we first train a high-dimensional neural network explicitly incorporating both spatial and temporal features (SFNN) to forecast measles incidence over 1,452 cities and towns from 1944 to 1965 and assess forecast performance over a range of different forecast steps. We also employ explainability (XAI) methods to shed light on how the neural network reveals mechanistic relationships when making predictions.

Following this we turn our attention to integrative approaches that have the potential to simultaneously provide high forecasting performance and mechanistic interpretabililty. Specifically, we focus on the so-called physics-informed neural network (PINN) methods, a class of integrative neural networks that incorporate physics differential equations into the model fitting procedure [20, 21]. PINN methods are able to preserve high predictive performance while incorporating and inferring scientific parameters, and have only recently been extended from physics differential equations to infectious disease mechanistic equations [6, 22]. While previous pioneering work [6, 22–24] has demonstrated the ability of PINN methods to improve disease incidence forecasts, the applications have not focused on long-term prediction and inference of the transmission dynamics in the context of endemic childhood infections. We build a PINN model which integrates a machine learning model directly with a mechanistic S-I-R model, and is able to address these shortcomings by augmenting the measles transmission dynamics with reconstructed latent susceptible dynamics from the TSIR model.

Our results demonstrate that appropriately designed machine learning models can outmatch more traditional mechanistic modeling approaches with respect to forecasting accuracy while effectively uncovering mechanistic infectious disease dynamics, in both a post-hoc and an integrative fashion. First, the high-dimensional neural network (SFNN), overall, outperforms the TSIR model for all forecast windows and in the majority of towns and cities in E&W, but with the most notable improvement for long-term predictions. The explainability (XAI) methods applied to our SFNN uncover the mechanism of hierarchical spread from large core cities to less populous towns without imposing such a mechanism *a priori*. Specifically, our results suggest that the relative role of spatial hierarchical spread increases as the population size of towns decrease, which is consistent with previous findings leveraging gravity model formulations [25]. Second, we compare the performance of a PINN model augmented with TSIR-reconstructed latent susceptible dynamics (referred to as TSIR-PINN) to a PINN model with naively constrained susceptible dynamics (referred to as Naive-PINN). We demonstrate that inclusion of the TSIR-reconstructed susceptible dynamics (in the TSIR-PINN model) improves the inference of disease parameters while simultaneously providing high forecasting accuracy. Together these findings illustrate the potential for a new suite of methods to provide improved integration between mechanistic models and machine learning approaches for infectious disease modeling, achieving high predictive performance while simultaneously ensuring accurate scientific inference of the spatiotemporal dynamics of measles and similar infectious disease systems.

## Results

### Neural network model (SFNN) outperforms TSIR model in forecasting endemic measles dynamics

The TSIR model estimates measles dynamics by leveraging incidence data and birth data [13, 18, 26] (see Materials and Methods for full model specification). It provides a computationally inexpensive and highly tractable alternative approach to the continuous-time S-I-R model. It has been shown to excel in short-term forecasting for measles incidence in large populous cities [17]. However, the TSIR model generally does not perform well for long-term forecasts. Furthermore, incorporating spatial interaction among multiple locations into the TSIR is a steep statistical challenge [27], which limits its utility for characterizing and forecasting (typically less regular outbreaks) in less populous towns whose population size less than the CCS.

With these deficiencies in mind we employ a neural network explicitly incorporating spatial and temporal features (SFNN). Specifically, our SFNN considers not only measles incidence lags as features, but also potentially important spatial features including the measles incidence lags in, and distances to, the nearest (ten) towns/cities and the (seven) highest population cities (see Materials and Methods for full model specification). The seven highest population cities were chosen because these were identified as having populations greater than that of the critical community size (CCS) of 300,000, an empirical threshold at which chains of infections are locally sustained [28].

Our results show that our spatiotemporally featured neural network (Figure 1) generally outperforms the TSIR for both short- and long-term predictions, across different population sizes (Figure 2). For very short-term predictions (e.g., when *k* = 1, where *k* is the number of biweeks ahead of the targeted prediction), our neural network model SFNN notably outperforms the TSIR in less populous towns where the TSIR model traditionally has struggled with. As the population size of the prediction target increases, the performance of SFNN gradually degrades (Figures 2B). As the forecasting time window widens (e.g., *k >* 4), the added predictive accuracy of the SFNN, in comparison with the TSIR, becomes more significant, but with an opposite trend: the performance of the SFNN now improves with the population size.

**Fig 1.**
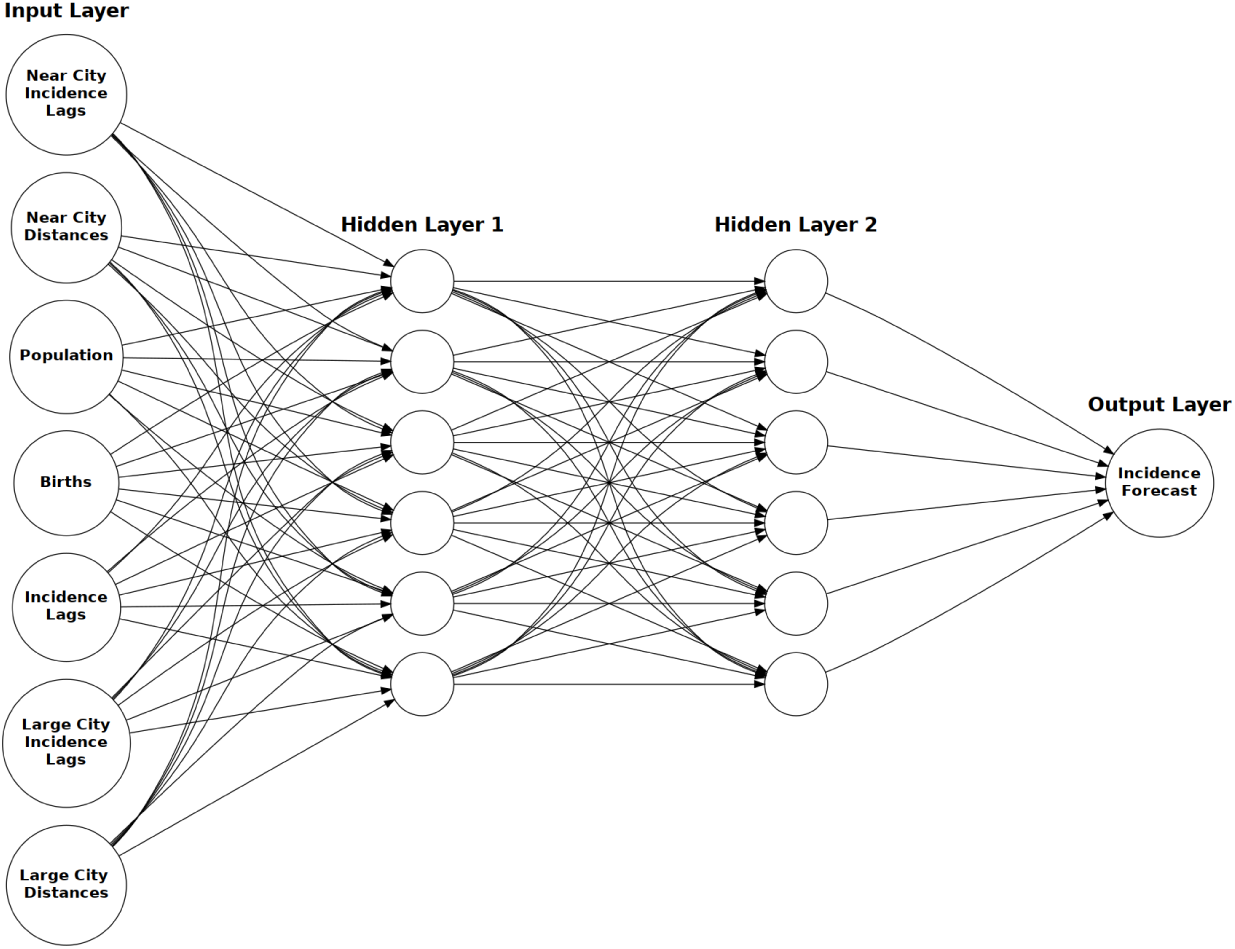
SFNN architecture. The SFNN architecture with grouped input features (grouped according to feature type), 2 hidden layers of dimension 962, and 1 output layer of dimension 1 for incidence forecasts.

**Fig 2.**
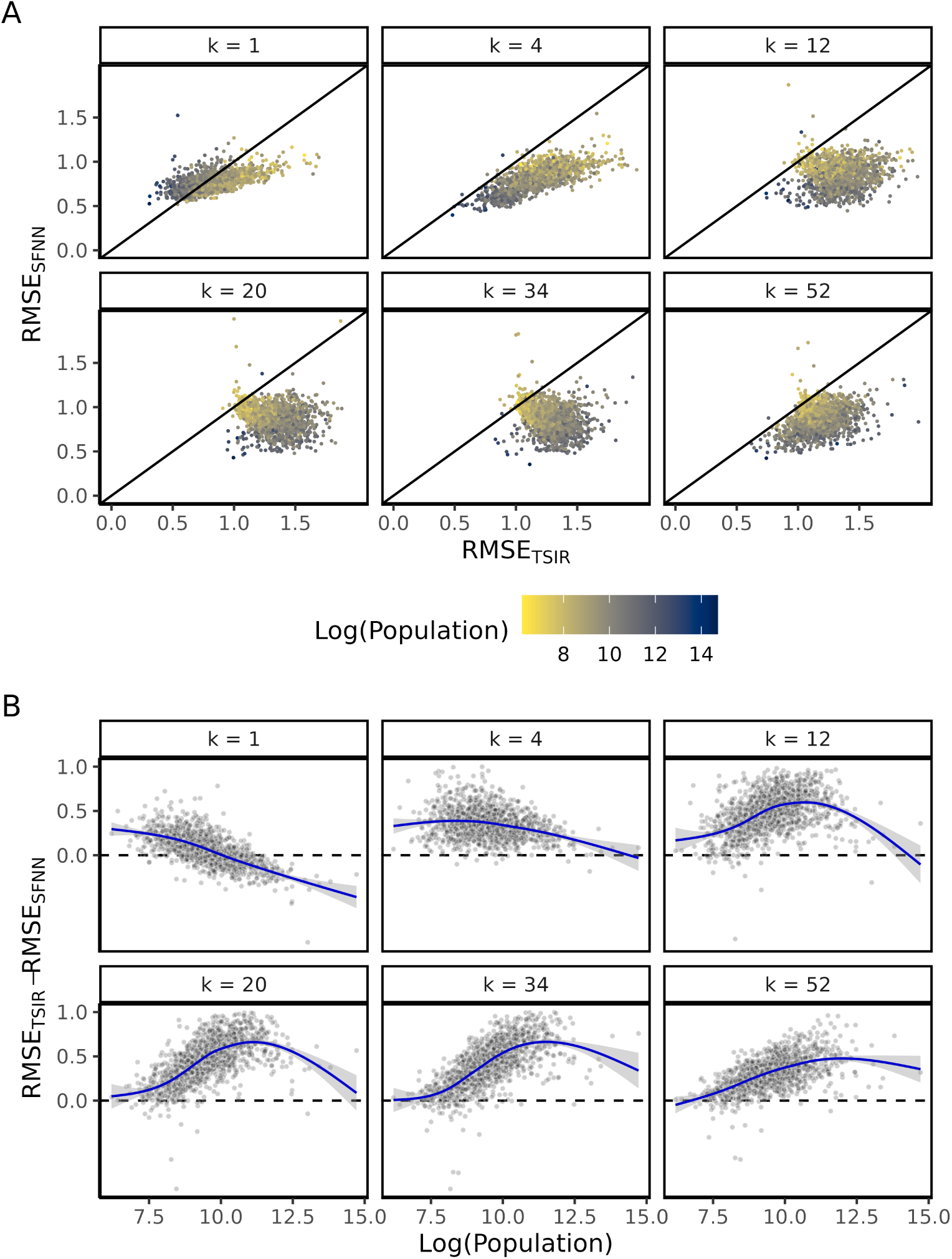
SFNN vs. TSIR model performance measured by Root Mean Squared Error (RMSE) of within-city-standardized log(incidence + 1). (A) Within-city SFNN RMSE versus TSIR RMSE, colored by log(population), faceted by *k*-step ahead forecast. (B) Difference between the within-city-standardized RMSE for TSIR and the within-city-standardized RMSE for SFNN; loess regression curves are fitted.

We also test whether our SFNN can capture annual to biennial bifurcation of measles epidemics in E&W caused by susceptible response to the late 1940s baby boom [18]. While our results (Figure S1) suggest that our SFNN trained on the limited data prior to 1948 has limited medium- and long-term predictive accuracy for the outbreak size, it largely captures the bifurcation of the seasonal pattern for smaller forecasting windows (*k ≤* 4).

### Neural network model (SFNN) can uncover mechanism of spatial hierarchical spread

Previous work has demonstrated the presence of gravity-like dynamics in measles outbreaks [16, 25, 29]. For instance, dynamics in small towns are shown to be driven by the mechanism of spatial hierarchical spread in which infections in large cities can serve as reservoirs for seeding infections in less populous regions [25]. To assess if the neural network is learning such a mechanism, we employ feature importance methods that estimate how predictions rely on information from certain features and groups of features. We specifically use SHAP values [30] to investigate the relative importance of a core city to the measles spread in locations with different population sizes (see Materials and Methods for more details).

Our results (Figure 3) show that the (lagged) incidence in large cities are relatively more important for less populous cities/towns. This suggests that our neural network model is able to reveal the mechanism of spatial hierarchical spread in the endemic measles spatio-temporal dynamics. This is notable both for the indication that our neural network SFNN is able to employ spatial features in a complex manner that reveals mechanistic dynamics, without explicitly imposing spatial hierarchy in the model *a priori*, and as an example of a post-hoc XAI method that reaffirms a theorized dynamic in a disease system.

**Fig 3.**
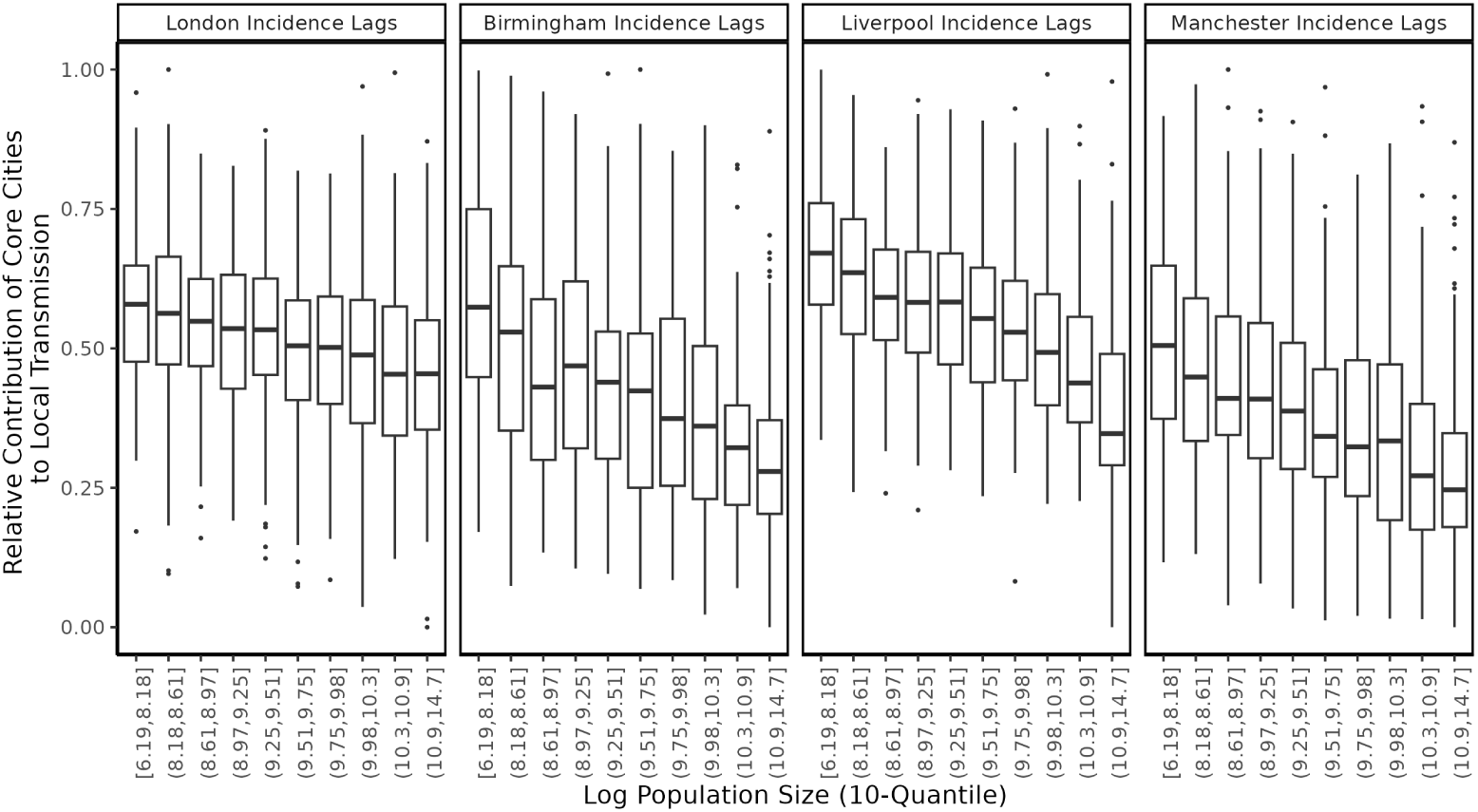
SHAP values uncover mechanism of spatial hierarchical spread. The SHAP value measures the relative importance of the incidence of a core city (e.g., London) for making incidence prediction among cities/towns with different population sizes, which can be heuristically treated as the relative importance to the local transmission of measles in a particular city/town. Core city incidence lag features are shown to be more important when predicting incidence for less populous cities/towns. Specifically, the mean relative absolute SHAP value for each of the core city incidence lag features has an inverse relationship with log population. Cities and towns are categorized (on the x-axis) into 10 groups according to the quantiles of their population sizes.

### Latent susceptible dynamics reconstruction using TSIR improves inference and forecasts of the integrative PINN framework

Next we turn our attention to integrative approaches that combine mechanistic and machine learning models. We consider the general conceptual framework of Physics-Informed Neural Network (PINN), a class of integrative neural networks that incorporate physics differential equations [20]. PINNs regularize a neural network by including a loss term which matches differential equations with observed gradient approximations garnered during the fitting process (typically using automatic differentiation methods). They hold the promise of preserving the high predictive capabilities and expressibility of neural networks while integrating scientific relationships directly into the model. Though PINNs are classically employed as a surrogate model for computationally intensive differential equation solvers [20], they also enable parameter inference and let (physics) dynamics partially drive predictions in an integrative fashion. These latter aims have been the primary impetus of existing methods to extend PINNs to spread of infectious disease [22] and are also the motive for us improving the PINN framework.

Here, we investigate the utility of a customized PINN model augmented by the reconstruction of the latent susceptible dynamics leveraging the TSIR model, referred to as TSIR-PINN (see Materials and Methods for full model specification). We compare the added benefit of our approach to a naive PINN model without such augmentation of the latent dynamics (referred to as Naive-PINN). We apply both models to London measles incidence data, and assess a two-year-ahead forecast window. Figures 4 shows that the Naive-PINN fails to make accurate predictions and parameter inference, while our TSIR-PINN model utilizing TSIR-reconstructed susceptible dynamics are able to capture and predict the transmission dynamics reasonably accurately. In particular, the TSIR-PINN model estimates an *R*_0_ value (Figure 4) which is largely consistent with previous estimates [18]. The TSIR-PINN model also outperforms the Naive-PINN with respect to test-set Mean Absolute Error (MAE) and correlation (Table 1).

**Fig 4.**
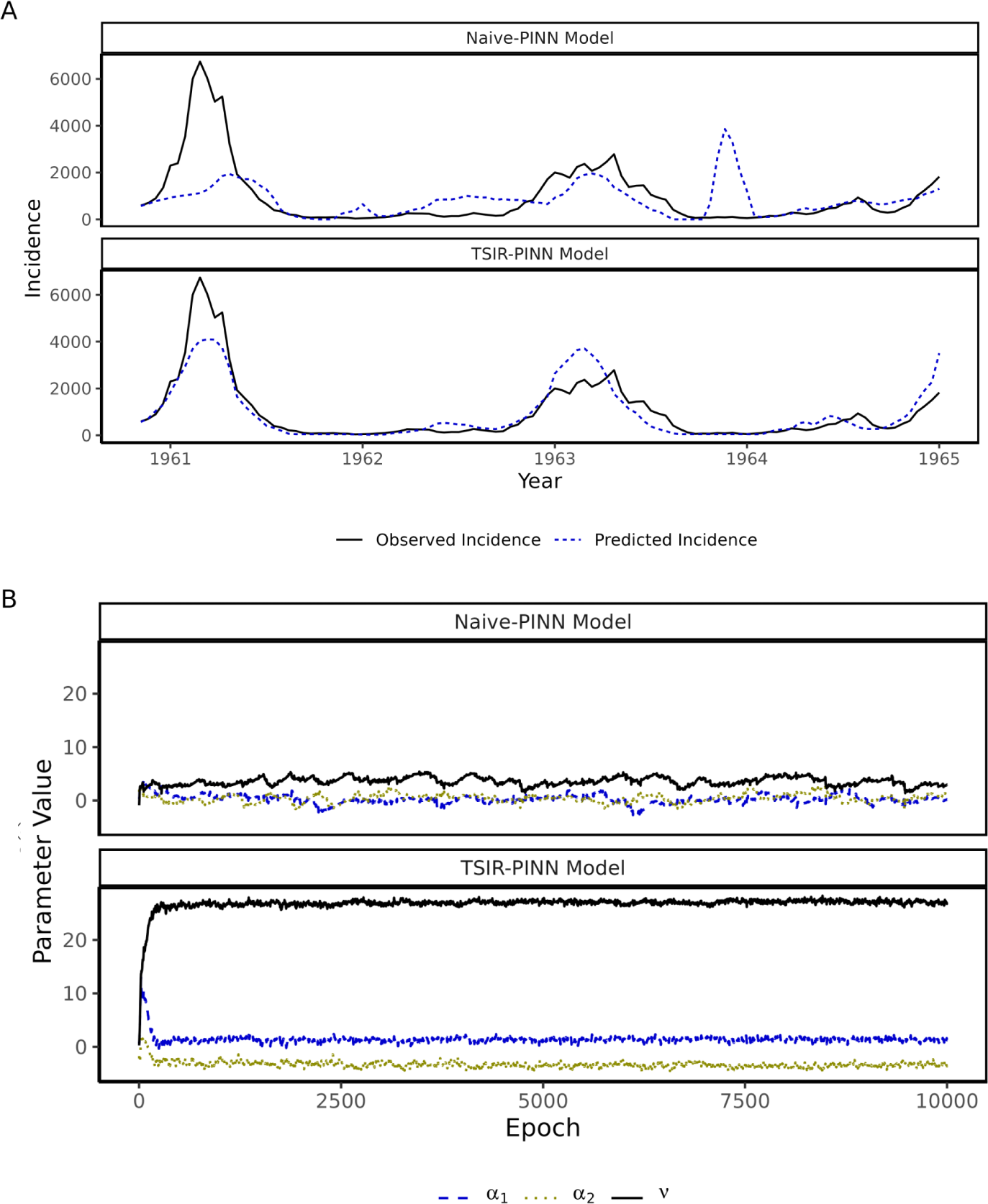
Test-set 52-step-ahead incidence predictions over time (A) and Inference of seasonal transmission rate (B) in London. (A) PINN test-set 52-step-ahead incidence predictions for TSIR-PINN and Naive-PINN models. (B) PINN parameter values are notably different between TSIR-PINN and Naive-PINN models over 10,000 epochs. The parameter *v* (black lines) correspond to the *R*_0_. Convergence is rapidly achieved when fitting the TSIR-PINN model, while convergence is less clear for the Naive-PINN model. More importantly, the TSIR-PINN model estimates an *R*_0_ of 26.9 which is broadly consistent with the literature, while the Naive-PINN estimates an *R*_0_ of 3.0.

**Table 1.**
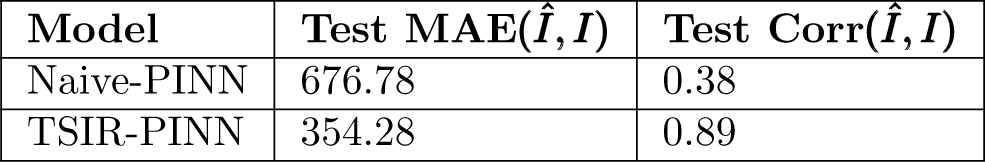
Performance of TSIR-PINN and Naive-PINN measured by test-MAE and test-correlation.

Our results suggest that including the TSIR-reconstructed (latent) susceptible dynamics (in our TSIR-PINN) can improve parameter inference while maintaining the predictive capabilities of a PINN modeling framework. These results provide important insights into more rigorously incorporating partially observed epidemic data into a PINN model, which may facilitate future developments and applications of PINN-based epidemic models.

## Discussion

Measles is among one of the most well-documented infectious disease systems and is known for its complex spatio-temporal dynamics. Spatiotemporal dynamics of measles infection, driven by interplay between seasonal forcing and susceptible recruitment dynamics [17], range from simple limit cycles to chaos, with the domination of stochastic extinction in small, highly vaccinated populations [18, 31, 32]. As such, measles serves as an excellent test bed for developing modeling techniques aimed at understanding similar nonlinear infectious disease systems.

Flexible machine learning approaches hold much potential for forecasting measles dynamics. However interpretation and inference is often difficult due to high dimensional model parameterizations and lack of scientific knowledge integration. Two broad classes of methods are suitable for improving mechanistic interpretability of machine learning models for infectious disease dynamics: post-hoc explainability (XAI) methods which conduct post-hoc analysis on model outputs to understand underlying drivers of predictions, and direct integration of mechanistic models or other scientific priors into machine learning models. Here we detail one example of each of these classes and demonstrate their effectiveness in accurately characterizing measles spatio-temporal dynamics while preserving high forecasting performance.

Our high-dimensional feed-forward SFNN overall performs well for all forecasting windows and the majority of cities. More noteworthy is its ability to outperform TSIR for the difficult forecasting scenarios of long forecast windows (ranging from six months to two years) and less populous towns with sparse, less regular outbreaks. Neural networks are known as a “black box” method, indicating that the way in which the model uses specific covariates to arrive at a forecast is not readily apparent from parameter inspection. This is the primary downside of employing such machine learning methods when compared to mechanistic and semi-mechanistic methods such as the TSIR, which provide ample opportunities for parameter inference and assessment in relation to scientific knowledge and hypotheses. To surmount this limitation there are a collection of post-hoc methods that allow methodical interrogation of machine learning output.

Our application of one such method, the SHAP value XAI calculation [30], is able to provide insights into how our SFNN predictions are being driven by a combination of input variables that has scientifically meaningful interpretation. Specifically we show that our SFNN uncovers the mechanism that outbreaks in large cities may influence measles transmission in smaller towns/cities. This is consistent with previously theorized mechanism which suggest a similar dynamic of hierarchical spread of infections from large cities to smaller towns [14].

While this post-hoc method is insightful and relatively straightforward to apply due to its lack of interference with model-training, we push the neural-network inferential capability further with a fully integrative PINN based model (TSIR-PINN) that incorporates reconstructed latent susceptible dynamics from the seminal semi-mechanistic TSIR model. By fusing mechanistic compartmental models with the neural network during model training, we are able to inspect disease dynamic parameter estimates while maintaining forecast performance. We show that by including the reconstructed latent susceptible population in our TSIR-PINN, both forecasting performance and parameter estimation are improved when compared to the Naive-PINN model (which does not utilize the augmented latent susceptible dynamics using TSIR). While PINN-based models have previously been applied to infectious disease data, our work is a step forward in terms of more rigorous inference of and integration with latent aspects of the transmission dynamics, which is crucial in enabling long-term forecast windows and deep mechanistic interpretability. Our results provide key insights into rigorously incorporating partially observed epidemic data into a PINN-based modeling framework, which may facilitate future developments and applications of PINN-based epidemic models.

There are several potential future directions we can explore. The PINN formulation introduced here provides solely point estimates of disease dynamic parameters, and one area ripe for further development is the incorporation of rigorous statistical uncertainty quantification into these methods that might enable probabilistic statements about both model parameters and predictive output. There is also potential for this work to be extended to other disease systems, incorporating mechanisms and assessing hypotheses that are specific to these areas of study. Furthermore, we have only applied the TSIR-PINN model to London measles incidence data, though there is potential to extend the TSIR-PINN to multiple cities (or all cities, such as with the SFNN) by explicitly incorporating gravity model mechanisms. Also, we have focused on comparing the relative performance of the TSIR-PINN to the Naive-PINN, and thus there remains room to further explore model formulations that may result in even higher prediction accuracy, such as shared embeddings or explicitly spatial or temporal architectures such Convolutional or Long Short-Term Memory Neural Networks (CNNs, LSTMs respectively) [33, 34]. Finally, there is potential for application of machine learning methodology that instead of imposing compartmental model structures *a priori* and inferring parameter values, focuses on hypothesis generation and model structure discovery in the context of infectious disease [35–38]. This could automate some of the fine-tuning of model structure required for these highly bespoke models and aid modellers at earlier stages in their research.

In summary, our results show that appropriately designed neural network-based models can outperform traditional mechanistic models in forecasting, while simultaneously providing mechanistic interpretability. Our work also offers valuable insights into the more effectively integrating machine learning models with mechanistic models to enhance public health responses to measles and similar infectious disease systems.

## Materials and methods

### Study Data

We train and assess our models on biweekly measles incidence counts across 1,452 cities/towns in England & Wales during the pre-vaccination period from 1944 to 1965 (Fig 5). Separate models are fitted for different *k*-step ahead forecasts, ranging from 1 to 52 biweekly time steps ahead.

**Fig 5.**
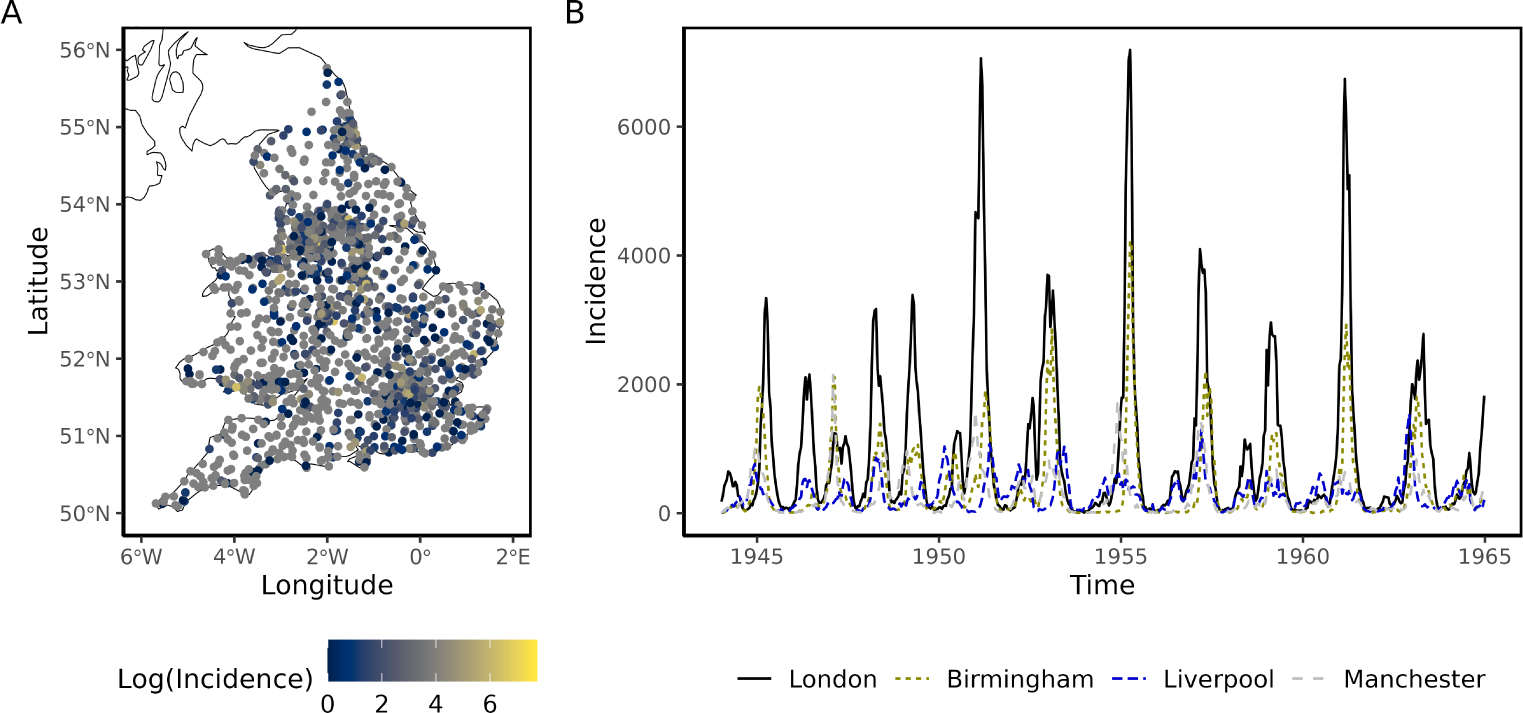
Measles cases in England and Wales. (A) Cities/towns colored by log measles incidence on first biweek of 1961. (B) Seasonal measles trend apparent across the four most populous cities from 1944 to 1965.

### Feed-Forward Spatial Feature Neural Network Model (SFFN)

For each *k*-step ahead we fit a separate feed-forward spatial feature neural network (SFNN) with 2 hidden linear layers of dimension 962, linear input/output layers, and ReLu [39] activation functions (Figure 1).

We include a range of features, including birth counts, population size, lagged incidence counts and lagged incidence counts and distances for the seven cities with a population higher than the critical community size of 300,000 which has previously been identified as “core cities” that drive epidemics in connected cities/towns. [28]. We also incorporate spatial features, including the lagged incidence counts of the nearest ten cities and their distances. This potentially enables the neural network to learn spatial dynamics that the TSIR model does not capture. Birth and population features are from the nearest time step less than or equal to *t − k* while still sharing the same biweek of the year. Lagged features range from *t − k* to *t −* 130, where *t* is the target time step.

Neural networks are fitted in Pytorch [40] using the Adam [41] optimizer with Mean Squared Error (MSE) loss, and are trained on incidence data ranging from 1949 to 1961 with incidence data ranging from 1961 to 1965 held out for testing.

### Time-series SIR (TSIR) model

We compare the neural networks to the TSIR (time-series susceptible-infected-recovered) model, a popular semi-mechanistic technique that has been shown to accurately capture the dynamics of measles outbreaks in major cities [13]. TSIR provides a computationally inexpensive and highly tractable alternative to the classic SIR compartmental model, and is described by the following equations:

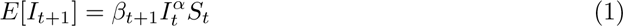

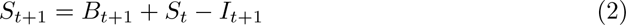

where *S_t_* is reconstructed as 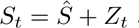 at each time step and with 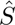 the average number of susceptible individuals in the population. *Z_t_* is estimated from equation 2 by regressing the cumulative births against the cumulative incidence as follows,

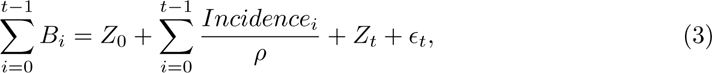

and the log-linearized equation 1.

For each *k*-step ahead, target time set *t*, and city, a separate TSIR model is fit on time steps *t −* 130 to *t − k*. One-step ahead forecasts are then made recursively with equations 1 and 2 until time *t* forecast is reached. We employ the **tsiR R** package for TSIR model fitting, and refer the reader to the package documentation for details not specified here [26].

### Neural network interpretability methods (SHAP)

We use the SHAP (SHapley Additive exPlanations) method [30] to assess neural network feature importance, specifically relying on sampling-based approximation methods [42, 43] from the Captum [44] Python library. SHAP values are estimated by randomly permuting (input) feature groups, calculating the change in model output due to a particular permutation and finally averaging across all permutations. Features are grouped according to lag type; that is, incidence lags are grouped together, high-population-city incidence lags are grouped together, etc.

In our analysis, we first estimate the normalized absolute SHAP value associated with a particular feature group for each observation. We then, within a particular city or town, calculate the average of all the normalized absolute SHAP values associated with a particular feature group of interest, across all the observations of that city or town. Together these provide a measure of the relative importance of a particular feature group for the predictions made for a city/town.

### Physics-Informed-Neural-Network model

The neural network architecture for the PINN models is different from the previously described neural network, due to incorporation of compartmental S-I-R equations and parameters in the model’s loss function. We start with a Feed-Forward Neural Network with 2 hidden layers of dimension 128, linear input layer, and a 2 dimensional output layer for the TSIR-reconstructed susceptible (*S^T^ ^SIR^*) and observed incidence (*I*). GeLU [45] activation functions are used on the hidden layers and a softplus activation function is used on the output layer. Features include time, lagged incidence counts, and lagged TSIR-reconstructed susceptibles, with the time feature transformed with Gaussian Random Fourier feature mappings [46]. Neural networks are again fit in PyTorch using the Adam optimizer with the same train/test as previously, though here we employ a Mean Absolute Error (MAE) loss comprised of the following components:

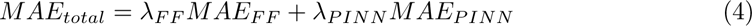

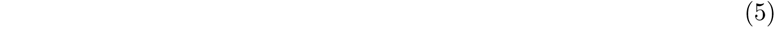

where *λ_F F_* and *λ_P INN_* are tunable hyperparameters, and

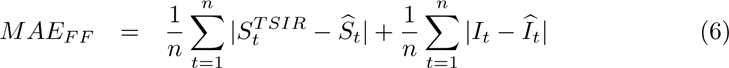

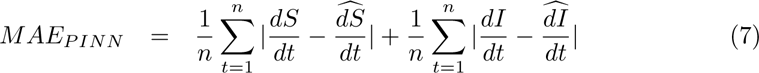

where 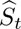 and 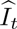 are the *FF* predictions at time *t*.

Here 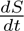 and 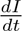 are the output of the following compartmental SIR equations at time *t*:

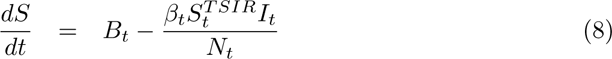

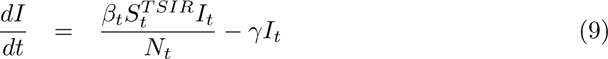

where *B_t_*is the number of births at time *t*, *β_t_*is a seasonal transmission rate at time *t*, *γ* is the recovery rate, *N_t_*is the population at time *t*, and 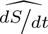 and 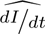 are the approximations of the relevant gradients, which are calculated at each epoch using *autograd* in PyTorch [40].

We parameterize *β_t_*as follows:

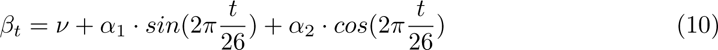

where eq. 10 implies a seasonal transmission rate with three free parameters: *ν*, *α*_1_, and *α*_2_. *ν* is the baseline transmission rate, while *α*_1_ and *α*_1_ are seasonal parameters controlling sinusoidal annual fluctuations.

We assume *γ* = 1 due to the measles recovery period being approximately equal to the biweekly scale of the data [47], thus the parameters employed in *β_t_* are the sole learnable parameters for this *MAE_P INN_* component of the loss. By matching 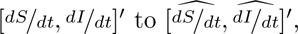 we are providing an unsupervised soft constraint on the neural network to adhere to compartmental equation dynamics and vice versa.

To assess the impact including the TSIR reconstructed susceptibles, we also fit versions of the above models with naively constrained latent susceptibles, such that all *S^T^ ^SIR^* components are replaced with *S^Naive^*, and are fit as unconstrained parameters.

## Data Availability

Measles data is available at https://github.com/msylau/measles_competing_risks

## Supporting Information

**Fig S1.**
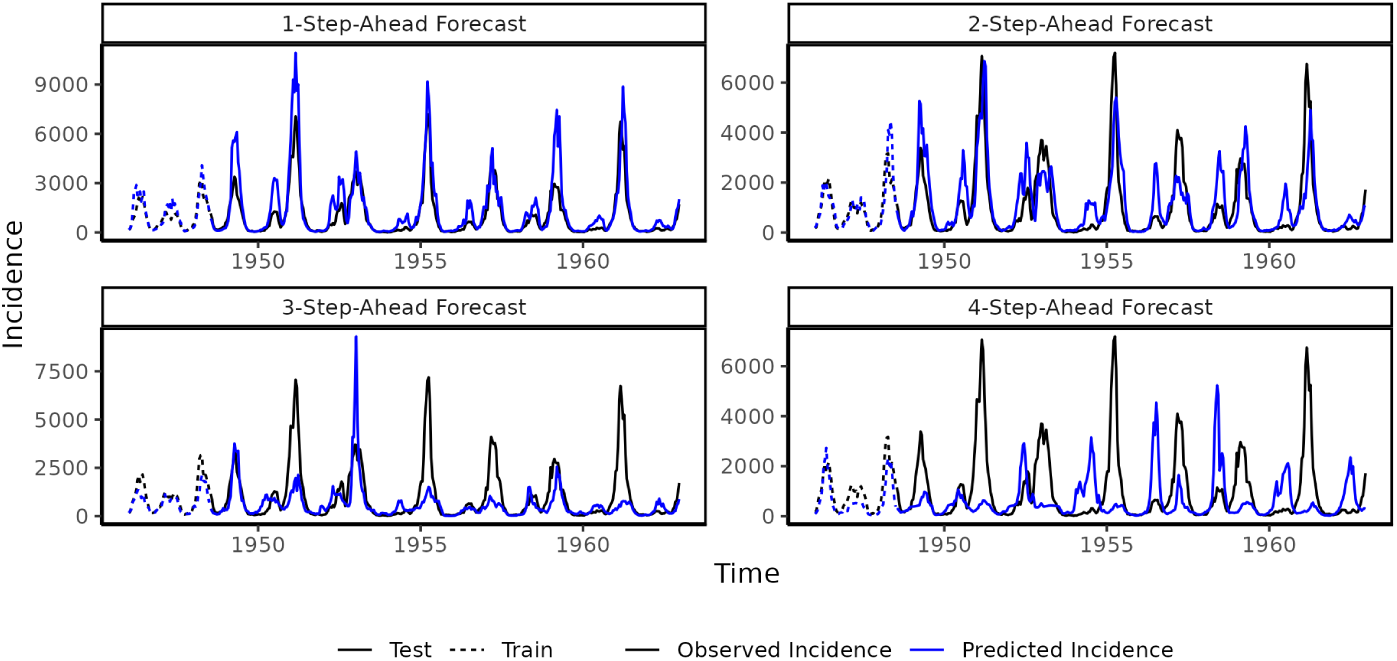
London SFNN bifurcation assessment. Our SFFN model trained on the limited data prior to 1948 predicts change of seasonality (i.e., annual to biennial bifurcation in late 1940s) in London, for steps-ahead ranging from 1-4. It is noted that, due to the lack of training data in this case, our SFNN does not perform well in capturing the magnitude of the incidence in general.

